# A model showing the relative risk of viral aerosol infection from breathing and the benefit of wearing masks in different settings with implications for Covid-19

**DOI:** 10.1101/2020.04.28.20082990

**Authors:** Gerald D Barr

## Abstract

**Background:** Widespread use of masks in the general population is being used in many countries for control of Covid-19. There has been reluctance on the part of the WHO and some governments to recommend this.

**Methodology:** A basic model has been constructed to show the relative risk of aerosol from normal breathing in various situations together with the relative benefit from use of different masks.

**Results:** The benefit from mask use between individuals is multiplicative not additive and although social distancing at 2 meters appears beneficial with regards to aerosol infectivity, in confined areas this is time limited requiring additional measures such as masks. The model shows the relative benefit of masks when social distancing is not possible at all times, or when in confined areas which can also be aided by efficient ventilation. Where a person is in one place for a prolonged period there is more risk requiring protection.

**Conclusions:** Masks should be used in the above situations especially at an early stage of an outbreak. Public health planning requires stockpiling of masks and encouraging everyone to have suitable masks in their household when supplies are normalised. In the absence of widely available good quality masks the use of a cloth mask will be better than no protection at all.

## Introduction

The WHO advised for Covid-19 from January 2020 that masks are not required for use by the general population, except for those with respiratory symptoms. [1] In April this changed to medical masks should be reserved for health care professionals [2], citing that mask wearing by the public might lead to complacency. However, it could be argued that informing the public that masks are not needed is potentially more damaging leading some individuals to believe that Covid-19 is only contracted by poor hand hygiene. The advice appears to be contradictory to the 2019 document for influenza which states that the approach should be multifaceted including, ‘Face masks worn by asymptomatic people are conditionally recommended in severe epidemics or pandemics, to reduce transmission in the community,’ although the publication does mention the possibility of reduced supply to health professionals and increase in prices as a consequence. [3] Even basic masks can help to prevent droplet spread which the WHO states is the primary spread of Covid-19 together with touching fomites, while acknowledging aerosol spread is an important consideration in hospitals.[4] Given the relatively high R number [5] and findings of aerosol spread in other respiratory viruses such as SARS and influenza this route must be a consideration in the community unless proven otherwise. The UK has adopted a similar policy that a decision cannot be made regarding general use of masks in the population based on having an insufficient evidence base. A rapid Cochrane review commissioned by the WHO assessing quarantine alone or with other public health measures to control Covid-19 does not mention masks. [6] A past Cochrane review [7] for influenza showed low level evidence in favour of general mask wearing but due to the lack of good quality studies could not make a positive recommendation pointing to compliance issues, but these studies do not translate to a pandemic with significant morbidity. A further review by Public Health England concluded that low level evidence suggested there was a benefit of masks for health care workers and also in the community, but community use could not be recommended. [8 ] It seems illogical in a pandemic with significant morbidity that only health care workers need protected when the driving force for their need is the numbers of people from the community being admitted to hospital. Greenhalgh referred to the precautionary principle when there is no evidence for doing something but where the risk of doing nothing could cause considerable harm in relation to recommending the use of masks in the general population [9] and Cowling asked for a rational approach to the use of masks. [ 10]

Aerosol spread occurs much more from coughing and sneezing, compared to speaking or normal breathing, however, some individuals produce excessively high amounts of aerosol particles > 10,000 per litre in normal breathing [11] which may be one mechanism for superspreading. From an observation in influenza [12] causing 100 per cent infection in a confined space it was postulated that for airborne infection there must be a relationship between the amount of virus expelled into the air, the ventilation of the area and the time exposed. The objective of this study was to construct a simple mathematical model to show the relative benefit of mask wearing in relation to the infectious dose of a virus in aerosol. A Health and Safety review found a six times reduction in bioaerosol using a surgical mask with an estimated 100 times reduction for an FFP3 mask, with much of the inefficiency of surgical masks being due to leakage or poor fit. [13] Cloth masks are acknowledged to give some protection which may only have an improvement factor of two.[ 14]

## Method

The model was formulated, based on the formulae described below, on an Excel spread sheet which allows input values to be changed easily and is available for general use on request. Input values available from the literature were used.

Infectious Dose *ID = P*(*av*) *EvF_1_* (*Bv /Ev*) *F_2_ t* where t is time in minutes. (Bv cannot exceed Ev i.e. max for *Bv/Ev* is 1)

Infectious dose (ID) is the amount of virus needed to be inhaled to cause infection. Many factors could affect this, and the bioavailability of the virus once inhaled together with mucosal defences. A review by Nikitin suggested a figure of 1.9 x 10^3^ for influenza. [15] Regarding fine aerosol that can penetrate more deeply into the lungs it is quite possible that a much lower dose is required. For this comparative study an ID at 1000 was used especially as this fitted best with scenario 2 (Table 2) and the original observation. Not all aerosol particles contain virus which may only be 10% of aerosol but can be more.[16] For this study 30% of particles being infectious was taken at av = 0.3.

For an infected person breathing out aerosol particles P = 500 / liter is used which is the cut off value between a low and high producer of aerosol.[11]

Ev is the volume of air expired by the infectious person per minute [tidal volume x breaths/min] for this study taken as normal breathing 500ml x12 breaths per minute = 6 litres /min.

Bv is the volume of air inspired by a non-infected person per minute [tidal volume x breaths/min ] for this study taken as normal breathing 500ml x12 = 6 litres /min. F_1_ is the expiratory filtration factor of a mask for an infectious person breathing out, F_2_ is the filtration factor for non-infected person breathing in. For no mask F is 1, for x 6 reduction is 0.167, x100 reduction 0.01. Using this data and that from a study [17 ] on the reduction factor, inspiratory and expiratory at 30, 50 and 80 l/min extrapolating to 6 1/min: cloth mask F_1_ is 0.75 F_2_ 0.5, surgical mask F_1_ is 0.333 F_2_ is 0.167, FFP3 F_1_ is 0.333 F_2_ 0.01.

At a distance under one meter the volume is not applicable due to risk of directly breathing the same air e.g. one person breathes in as the other breathes out. Greater than 1 meter the exhaled infectious air is dispersed into a volume V.

Ve is the amount of ventilation, given by the amount of original air remaining after replacement with fresh air per minute.

The time evolution of the infectious particle concentration *C* in the volume *V* is given by:

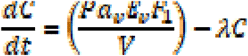

The time evolution of the inspired dose *N* is given by:

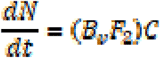

For zero decay constant *λ* the solutions are (where we consider that the infectious person has been present for a time *T_start_* before the uninfected person arrives):

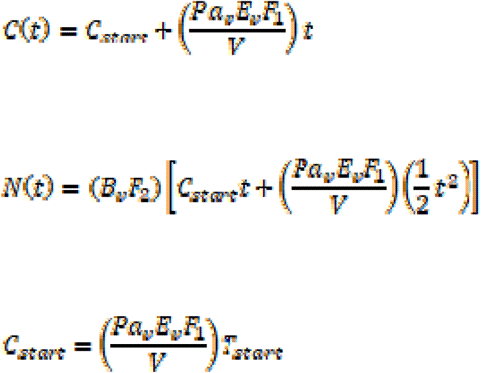

For nonzero *λ* the solutions are (where we allow a different decay constant *λ_pre_* during the warm-up phase before the uninfected person arrives):

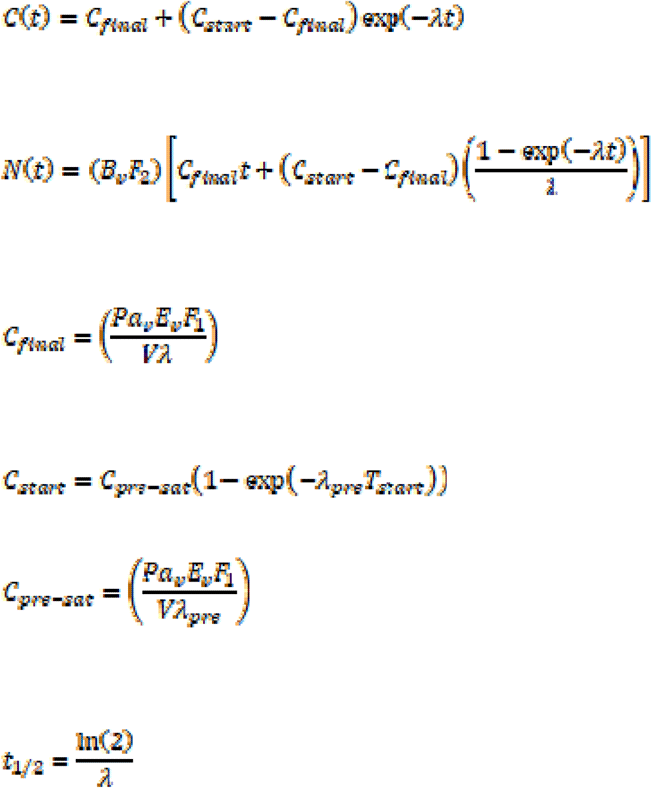

The aerosol absorbed by each person inhaling would reduce the number of infectious particles also increased by use of a mask affecting exhalation too. Fx is the inhalation filtration factor for the infectious person and Fy is the filtration factor for the non-infected person exhaling. Using the calculated loss from a medical aerosol study of absorption giving an estimated 45% of particles absorbed. [18]

The absorption loss during inhalation is given by:

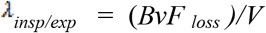

Where *F_loss_* is the effective fraction of particles lost in one complete breath and accounts for the effect of any masks during breathing in or out. The contributions of multiple individuals could be added to this value.

*F_abs_* is the fraction of particles absorbed with no mask then the effect of a mask can be estimated as:

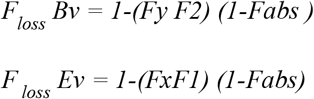

The exponential decay constant *λ* represents all loss processes which can be combined as:

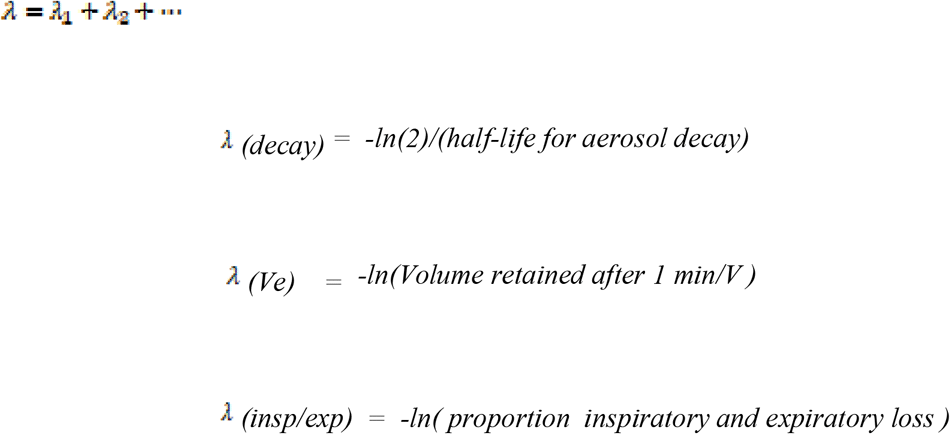

## General comments and limitation

This is a comparative analysis not an actual situation for any particular virus although the values used from the literature will give an approximation for Covid-19. Any of the chosen parameters could be less than the actual for instance the aerosol may only have 10% infectious particles, ID could be 3000 but equally particles exhaled by an individual could be 10 times more. The mask, no mask and ventilation analysis, however, is consistent and relative to these values.

The model assumes that the particles expired are instantly diluted to give a uniform concentration in the volume. In reality the distribution in space would be complex and so *V* must be regarded as an effective interaction volume rather than the precise volume of any particular region. Additionally, ventilation and convection or diffusion would create a complex spatial and temporal dependence of the concentration so the contribution to A by these processes must be considered as an effective value to model the loss of the virus from the system and not a precise value. It does not take account of localised directional air flow which in theory could increase the infectious dose i.e. directing higher concentration aerosol towards a person.

Decay of aerosol will vary for virus, particle size and humidity. Adams et al. found a sharp decay at the start for Reo virus, although this was considerably reduced by humidity, with recovery over longer periods [19] which may be due to smaller particles surviving longer. Cowling found that 87% of particles [20] emitted are less than 1 micron, at this size there is less potential space for a virus, for instance an aerosol particle 1 micron in diameter is much more likely to have a maximum of one SARS-CoVid-2 particle measuring 0.12 microns in diameter. Smaller particles can stay airborne for hours in a humid environment with only a 10% loss of infectivity. [21] Yang found enough influenza virus in the air to cause infection in different public places and that a steady state will be reached. [22] For this study it was assumed one infectious particle releases only one viral particle and for the calculations the median aerosol half-life of 1.2 hours for SARS-CoVid-2 found by van Doremalen was used. [23]

It is not certain what the minimum cumulative dose for infection is and over what period i.e. the minimum number of virus particles that nonspecific defences can cope with per hour or minute. It is assumed the ID is cumulative at least over a relatively short period for this study minutes to a few hours.

## Results

The model shows for two people, one infectious and the other non-infected, the protection from wearing masks is multiplicative not additive. Scenario 1 (table I) shows a high risk being close to someone breathing normally, potentially only for a few minutes. Wearing a mask will reduce the number of infectious particles which could mean reducing the chance of infection or even having a milder infection. Combining a surgical mask in the infectious and non-infected person gives a 2.8 times improvement from a surgical mask only on the non-infected person, and a 17 times improvement from no mask at all. The more people wearing higher quality masks will give more improvement and less risk of infection. Scenario 2 (table II) with two people on a half hour car journey reaching the ID by the end. There is a greatly reduced infectious dose with a non-infected person wearing an FFP3 mask, but all the masks reduce the ID. Both wearing a surgical mask is of considerable benefit although not as good as the non-infected person alone wearing a FFP3 mask. Scenario 3 (table III) shows that an infectious person in a confined area for 3 hours with no ventilation will give a high risk of infection. Such a situation could be a taxi where the driver is not separated from the passengers, or an infectious person in a cubicle with little ventilation. When someone enters the benefit of FFP3 is high but also there is a marked improvement by both wearing surgical masks. As there is only a short time benefit from a surgical mask alone or a cloth mask then ventilation with clean air is needed for further improvement. Figure1 shows in a restricted volume using cloth masks the difference with 5% of the air being replaced with clean air per minute. Scenario 4 (table IV) shows a person in an enclosed area with a larger volume for 3 hours and a non-infected person entering. This gives a low risk situation, but a cloth mask can give an even larger improvement. Scenario 5 (table V) shows two people socially distanced in a confined area such as a workplace. It shows how social distancing, which is based on how far droplets spread, can work for aerosol spread but it is time limited and needs to be extended with masks and ventilation over a longer period. Interestingly the effect of both wearing surgical masks was greater than 200 litres per minute of air replacement but both combined made an extremely large difference.

**Table I.** Scenario 1

|  |  |  |
| --- | --- | --- |
| Close by, breathing the exhaled air of an infectious person with no dilution, $av$ is taken at 0.3, both breathing normally so $E_v$ and $B_v$ are each 6 l/min | | |
| ID Infectious dose is 1000 particles |  |  |
| ID 1000 = $500 \times 0.3 \times 1 \times 6 \times (6/6) \times 1 \times 1 \times t$, $t = 1000/900$ **t = 1.1 mins** | | |
| Non infected wearing surgical mask ($F_2 = 0.167$) **t = 6.7 mins** | | |
| Both wearing surgical mask ($F_1 = 0.333$ $F_2 = 0.167$) **t = 19 mins** | | |
| Non infected surgical mask ($F_2 = 0.167$; Infected FFP3 mask $F_1 = 0.333$) **t = 19 mins** | | |
| Non infected FFP3 ($F_2 = 0.01$ ; infected surgical mask $F_1 = 0.333$) **t = 334 mins** | | |
| Both wearing cloth mask ($F_2 = 0.5$ $F_1 = 0.75$) **t = 2.96 mins** | | |

**Table II.** Scenario 2

|  |  |  |
| --- | --- | --- |
| 2 people in a car one infected, 2 m apart, Volume $2 \text{ m}^3$ no ventilation, $t = 30$ mins<br>Fresh uncontaminated air at the start | | |
| No masks | ID = 1078 p | t 30 mins |
| Non infected wearing surgical mask | ID = 177 p | t 30 mins |
| Non infected wearing FFP3 | ID = 11 p | t 30 mins |
| For both using surgical mask | ID = 58 p | t 30 mins |
| Both using cloth mask | ID = 396 p | t 30 mins |

**Table III.** Scenario 3

|  |  |  |
| --- | --- | --- |
| 2 people in confined space such as a car 2 m<sup>3</sup>, one infected person already present for 3 hours and no ventilation. |  |  |
| Starting concentration at 180 minutes is 35 p/l |  |  |
| No masks | ID is 1060 p | at 5 mins |
| Non infected FFP3 | ID is 1011 p | at 511mins |
| Non infected Surgical mask | ID is 1011 p | at 29 mins |
| Non infected Cloth mask | ID is 1056 | at 10 mins |
| Both wearing surgical masks from the start | ID is 1001 | at 95 mins |
| Relatively short time so ventilation with fresh air is needed unless using FFP3 or both using surgical masks |  |  |
| Introducing ventilation with 5 % of air replaced / min from the start of the three hours |  |  |
| Starting concentration at 180 minutes is 7 p/l |  |  |
| Non infected with no mask | ID is 1008 | at 24 mins |
| Non infected with cloth mask | ID is 1003 | at 48 mins |

**Figure 1.**
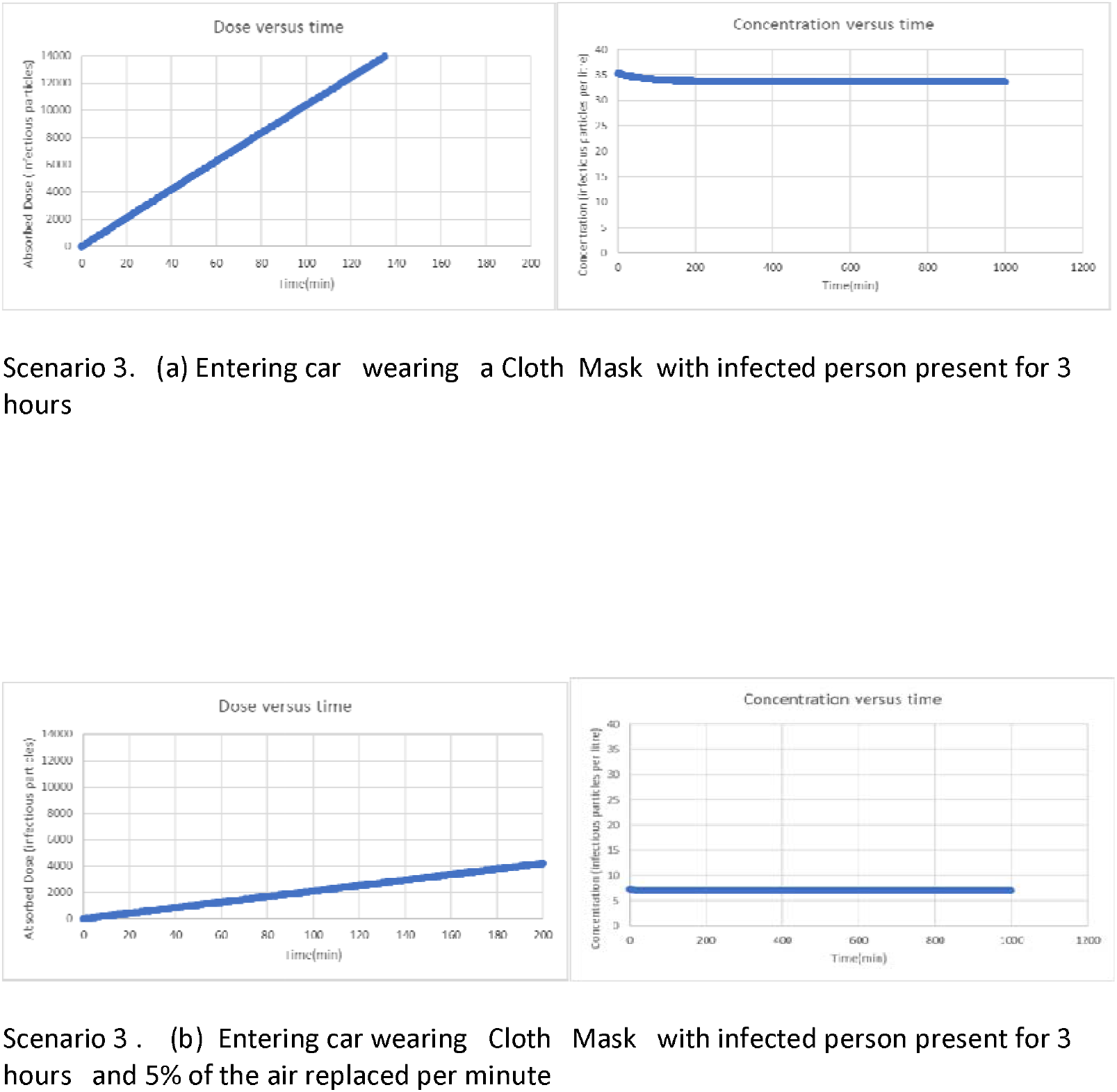
Scenario 3 Cloth Mask

**Table IV.** Scenario 4

|  |
| --- |
| Infected person in a room or working in a small shop 30 m<sup>3</sup> for 3 hours, no ventilation |
| Starting concentration at 180 minutes is 2.6 p/l |
| Non infected with no mask ID 1004 at 63 minutes |
| Non infected with cloth mask ID 1003 at 122 minutes |

**Table V.** Scenario 5

|  |
| --- |
| 2 people socially distanced at 2m in a confined area with a 4m<sup>3</sup> corridor of air between (assuming the air is static and no diffusion to a wider volume), one infectious and no ventilation, time to reach ID of 1000 |
| No masks ID of 1027 reached at 42 mins |
| Both wearing surgical masks ID of 1000 reached at 243 mins |
| With ventilation 5% air replaced per minute (200 litres/min) |
| No masks ID of 1003 reached at 62 minutes |
| Both wearing surgical masks ID of 1001 reached at 866 mins |

## Discussion

This study using mathematical modelling shows the relative benefit of wearing masks in different situations. Wearing a mask is beneficial at every aspect of infection modality as it can reduce aerosol inhalation with varying degrees of efficiency, from cloth to FFP3, and all masks will reduce large droplets. Reducing both will reduce fomite contamination; the wide distribution of virus in CoVid-19 infected patients’ rooms shows that aerosol deposition is important [24] in addition to droplets and direct touch. Any reduction in the ID could be significant by preventing infection or reducing viral load and degree of infection. [25] Given the possibility of anyone being infectious due to asymptomatic spreaders [26] high risk situations can be identified where greater protection is needed, especially where a person is in one place for a prolonged period or people are in close contact even for a short time, then use of masks is important. In addition to masks good ventilation and possibly air filtration could be important in workplaces or anywhere social distancing is time limited by a confined area. It should be noted that ventilation can be beneficial or harmful depending on air flow patterns [27] and input from health and safety and occupational health agencies would be useful in minimising risk in such places. Better quality protection is needed in environments such as public transport where airborne infections have been identified [28,29], inside shops where social distancing cannot be fully maintained at all times, or anywhere there is poor ventilation. The model suggests validation of strategies already in use such as putting a mask on a patient and ventilation of rooms by opening windows. Although social distancing at 2 meters appears to reduce infection risk from airborne virus this is time limited unless there is extremely good ventilation such as equivalent to being outside. If this is not the case and contact is maintained for more than a few minutes even with social distancing masks should be used. The better quality the masks the longer the contact time can be maintained with the same risk of infection or equally over the same time with a corresponding lower risk of infection.

Early use of masks in a pandemic is essential with good quality masks as part of a multifaceted strategy. Those returning from infected areas should use masks immediately together with isolation. Reductions in infections early on will have considerable benefit due to the number of people each infected person infects. Tracht et al produced a model for influenza that showed with 50% compliance in the population wearing N95 masks, there would be more than a 36% reduction in the number of cases. [30] They found less benefit for surgical masks but may have underestimated the combined efficiency of wearing surgical masks which this study shows a 17 times reduction in infection risk. Reducing the numbers of infected people early on will give contact tracing and isolation a much better chance to work. Models in pandemics should include aerosol spread and mask use [31], noting there was no mention of either in the models published by the UK government. [32,33] If there is an epidemic with rapid spread in a population with high morbidity a warning is sounding that in that country, or for other countries as soon as any spread out with that country occurs, plans for implementing the general use of masks should be engaged. The evidence for this is in the countries that have good control of the outbreak even with an initial exponential rise in cases, such as China, Japan, Korea, and Taiwan, although it is confounded with other measures such as contact tracing and social distancing. However, to suggest a deviation from an effective policy which includes the use of masks requires strong evidence that masks are of no benefit. Lockdown has been shown to be an effective measure against Covid-19 [34] but is not without cost. [35] Avoidance of lockdown in countries such as Taiwan or Hong Kong cannot be solely attributed to contact tracing or hand hygiene alone and addition of general mask use is recommended. [36] This can help keep numbers from rising so fast that contact tracing is overwhelmed as evident in the UK and USA. Random testing is useful for determining release from lock down [37] but with a greater than 1% mortality mitigation is not acceptable without imminent prospect of a vaccine and the aim needs to be suppression. Anytime lockdown is considered masks should already be in use. If use of masks shortens lockdown is by only a few days, it would be cost effective. This is illustrated in table VI which gives an approximate estimate for the UK noting the 3 month reduction in GDP also includes a period before lockdown. [38]

**Table VI.**
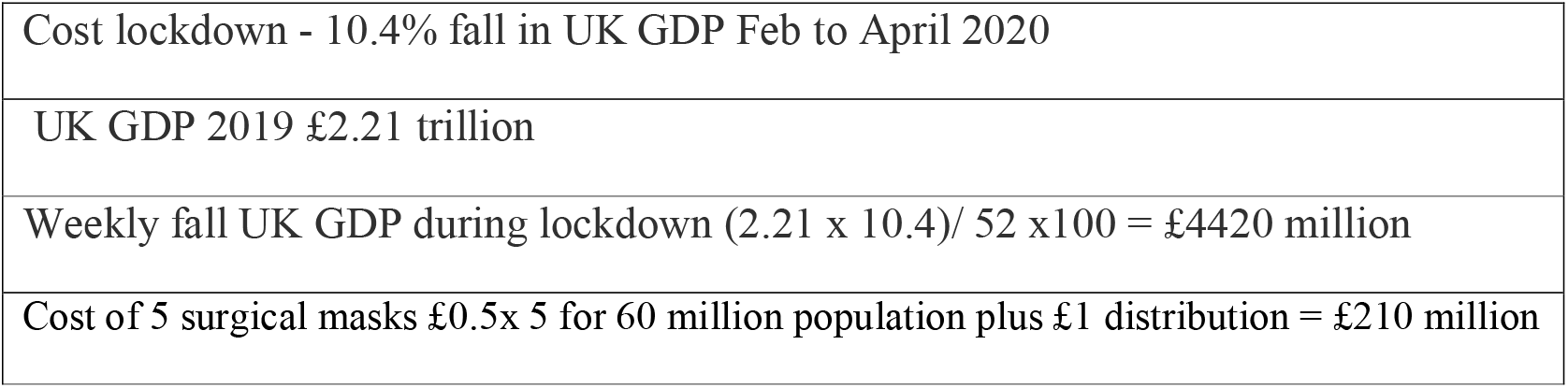
Approximate Cost of 1 week of UK Lockdown v Masks

|  |
| --- |
| Cost lockdown - 10.4% fall in UK GDP Feb to April 2020 |
| UK GDP 2019 £2.21 trillion |
| Weekly fall UK GDP during lockdown $(2.21 \times 10.4) / 52 \times 100 = £4420$ million |
| Cost of 5 surgical masks £0.5 x 5 for 60 million population plus £1 distribution = £210 million |

For Covid-19 the lack of resources is all too evident, but a lack of resources should not dictate public health policy, public health policy should ensure resources are available when needed. With regard to masks national stockpiles are necessary, high grade protection for the care sector and at least surgical masks for the general population. The more people that can use higher grade protection such as N95 especially on public transport and crowded places the better but if there is a national supply of surgical masks there would be a fall back for those unable to tolerate the higher grade masks or unable to procure them. Manufacturers increasing the shelf life from 5 years to 7 years would make this much more feasible. In addition to a national endeavour once supplies are normalized at a domestic level all households should be encouraged through public health information to keep a supply of good quality masks that can be used for dusty activities and DIY, then replaced when dirty. Alternatively, there are reusable masks such as sports masks which are usually available with PM2.5 2.5 micron pollen or dust filters but could be supplied with additional PM1 1 micron filters for pandemics, balancing comfort with reduced efficiency compared to N95 masks. Exhalation valves in theory are designed to reduce moisture and CO_2_ but it appears with the current design most valves do not work at low levels of breathing [39] which allows a protective effect in exhalation. Masks without exhalation valves still seem preferable for general use but in the future improving the exhalation filtering and fit to equal the efficiency of inhalation filtering would improve the combined efficacy of wearing masks considerably.

Compliance has been a problem in western countries even as far back as the Spanish Flu pandemic on the other hand masks have developed significantly since then. The health belief model [40] shows perception of risk of death and to health are important factors in compliance, also when livelihood becomes affected. In Hong Kong during the SARS epidemic the public mask usage was around 65%, but for asymptomatic individuals as low as 21.5% for H1N1 [41] rising to over 95% for Covid-19. A strong and consistent public health message is needed such as the campaign in Czechia, with visual media messages, “ I’m protecting you you’re protecting me,” also introducing the compulsory public wearing of masks early on. This compares to the USA and UK where for weeks even after the exponential rise in cases and hospitals being overwhelmed the official message was that masks were not needed. Not surprisingly this has to led to a relatively low compliance and the need for mandatory mask use on public transport and in shops including fines for those non-compliant in Scotland. Generally, it is acknowledged that older people and females [31] will engage more with mask wearing. Compliance within households will be low in the absence of infection [42] but the best option is to keep households free from infection by general population measures.

Most of the issues postulated that masks in the community would be detrimental are hand hygiene issues, such as touching the mask with unclean hands, meaning public educational information is needed and similarly advice on fitting can be addressed. Concerning other proposed disadvantages by Lazzarino [43], such as increase in risky behavior, there is no evidence in countries that have high mask usage. The argument is similar to the compulsory wearing of seat belts where there is no evidence of increased risky driving behavior. [44] In fact masks can be a reminder that other measures such as social distancing are in operation. There are some individuals with respiratory disease that would find using a mask difficult but the more people using masks will provide more protection for them and if necessary powered respirators are available. Lazzarino’s proposed increased respiratory rate secondary to CO2 retention from the mask causing increased exhalation of respiratory particles to those unable to wear mask, for instance with COPD, can be disproved by our model. Taking a person with COPD and no mask and respiratory rate of 12 liters/min, together with an infectious person wearing a FFP3 mask the respiratory rate would have to increase more than 4 times to be more infectious than normal breathing and no mask. A mask would not be tolerated for long if causing a fourfold increase in respiratory rate. One misquoted study shows cloth masks allow more infections compared to medical masks in the healthcare setting [45] and that cloth masks were worse than the control group but as most of the control group used medical masks there is no study showing that cloth masks are worse than no mask. As viruses cannot multiply on a mask there is no exposure situation where wearing a mask can be worse than not wearing one, although it does make sense to keep the mask clean and dry to maintain efficiency and prevent bacterial or fungal colonization. Protection against eye risk is not a case against masks, which is more likely when close to someone from droplets compared to aerosol (taking the study results and the surface area of the lungs compared to the eyes). In this situation a minimum of everyone using a basic mask and hand hygiene, similar to precautions needed in commercial food preparation, will help to reduce eye contamination risk. Higher efficiency masks can be more difficult to tolerate for long periods especially in humid environments, however, surgical masks are easy to wear for long periods and the combined effect of everyone wearing one giving a 17 times reduction in infection risk is not insignificant.

## Conclusion

The model shows the relative risk of aerosol in various situations together with the benefit from use of masks which is multiplicative. Social distancing at 2 meters appears validated but in confined areas is time limited and the use of masks in addition to efficient ventilation is important. Where social distancing is not possible at all times or anyone (a potentially infectious person) is in a confined area for a long time there is a higher risk of infection requiring protection. Masks should be used at an early stage as widespread use of efficient masks could have a large impact on control and spread of infection. Public health planning requires stockpiling of masks and encouraging everyone to have suitable masks in their household when supplies are normalized. In the absence of good quality masks, the use of a cloth mask is better than no protection at all.

## Data Availability

Spread sheet is available

## Acknowledgement

to Timothy Heelis for providing assistance with the numerical calculations.

## Notes

### Competing Interest Statement

The authors have declared no competing interest.

### Funding Statement

None

### Author Declarations

none this is a mathematical model and literature review no ethical approval is necessary .

